# Structured large language model extraction of clinical factors from electronic health record text supports scalable psychiatric severity prediction

**DOI:** 10.64898/2026.05.11.26352839

**Authors:** Callum Stephenson, Alessandra Camassa, Margot Wagner, Amirhossein Shirazi, Nazanin Alavi, Mohsen Omrani

**Affiliations:** OPTT Inc., Toronto, ON, Canada; Department of Psychiatry, Queen’s University, Kingston, ON, Canada; Salk Institute for Biological Studies, La Jolla, CA, USA; Institute for Neural Computation, University of California, San Diego, San Diego, CA, USA; Centre for Neuroscience Studies, Queen’s University, Kingston, ON, Canada; Kingston Health Sciences Centre, Kingston, ON, Canada; Centre for Digital Health Research, Providence Care Hospital, Kingston, ON, Canada

## Abstract

**Background:** Mental health systems face escalating demand that exceeds clinician capacity, making accurate severity-based triage a critical bottleneck. Severity assessment guides treatment intensity, resource allocation, and risk management, yet most clinically relevant information remains embedded in unstructured electronic health record (EHR) narratives, limiting its utility for scalable decision support.

**Objectives:** This study evaluates whether a single large language model (LLM) can autonomously extract clinical factors from psychiatric EHR narratives, derive predictive weights from those factors, and use the resulting structured representation to predict clinician-implied severity at scale.

**Methods:** From a Mayo Clinic repository of more than 2.7 million encounters, 15,000 de-identified psychiatric notes were sampled into a 5,000-patient discovery cohort and a 10,000-patient replication cohort. The same LLM (Llama 3 8B Instruct) extracted 17 background clinical factors and 3 treatment-action factors from each note. Severity reference labels were derived from the treatment-action factors using pre-specified clinical criteria. The LLM independently derived two factor-weight dictionaries from the discovery cohort: one capturing risk-oriented predictors of severe presentations and one capturing protective predictors. Five weighting conditions were then evaluated against the severity labels: the two LLM-derived dictionaries, two controls (LLM-derived variables with randomized weights; clinically irrelevant variables with arbitrary weights), and an unweighted zero-shot baseline. Performance was assessed across 928 valid iterations in the replication cohort.

**Results:** LLM-derived structured conditions significantly outperformed all controls and the baseline, with statistically equivalent performance between the two structured conditions. Improvements in precision and recall were balanced, indicating gains in discriminative capacity rather than threshold shifts. The variables and weights the LLM derived as predictors of severe presentations aligned closely with established clinical determinants of psychiatric severity.

**Conclusion:** A single LLM can derive clinically meaningful factor weights from unstructured EHR narratives and use them to predict psychiatric severity at scale, supporting a viable path toward interpretable, scalable triage in resource-constrained mental health systems.

## Background

Mental health systems are facing unprecedented demand that far exceeds available clinical capacity. The rising prevalence of depression, anxiety, trauma-related disorders, and suicidal crises has placed sustained pressure on care delivery models that rely on one-to-one, first-come, first-served clinician encounters. At the same time, the supply of trained mental health professionals, particularly psychiatrists, has not scaled to meet this demand. As a result, many patients experience prolonged wait times, delayed interventions, or misalignment between clinical need and the level of care they receive.

Critically, not all patients require the same intensity or type of mental health care. While some presentations warrant urgent psychiatric intervention, hospitalization, or intensive risk management, many others can be safely managed through outpatient follow-up, psychotherapy, or lower-intensity supports. The core challenge for health systems is therefore not simply expanding capacity, but accurately identifying which patients require the scarcest resources and which do not. In current practice, these triage decisions depend almost entirely on expert clinician judgment, which is difficult to scale and itself constitutes a major bottleneck in overstretched systems.

Despite the centrality of triage to mental health care delivery, there remains no rigorous, objective, and scalable method for evaluating individual patient severity using routinely collected clinical data. Accurate assessment of mental health severity remains a cornerstone of effective psychiatric care, influencing decisions related to resource allocation, treatment intensity, and risk management. However, symptom severity evaluation in routine practice is often inconsistent, context-dependent, and variably documented across providers and settings (Zimmerman et al., 2018). Although standardized instruments such as the Patient Health Questionnaire (PHQ-9) and the Generalized Anxiety Disorder-7 (GAD-7) provide reliable frameworks, they are not administered at every encounter and are rarely available at scale (Kroenke et al., 2001; Spitzer et al., 2006). Additionally, these measurement techniques do not consider information such as medical history, psychosocial factors, or comorbidities, which are essential for determining mental health severity and treatment recommendations. Consequently, much of this clinically relevant information needed to assess mental health severity and make treatment recommendations is embedded in unstructured narrative documentation within electronic health records (EHRs), limiting its utility for automated or population-level decision support (Kariotis et al., 2022). Hence the gap between structured survey data and unstructured EHR data.

Large language models (LLMs) can now analyze vast amounts of unstructured clinical text. Transformer-based architectures identify diagnostic patterns and extract clinical concepts with high fidelity (Crema et al., 2022). In mental health research, these models already detect suicidal ideation and analyze behavioural signals directly from narrative text (Stephenson et al., 2025; Xu et al., 2025). However, current methods rely on keyword heuristics or opaque end-to-end classifiers. These approaches lack the interpretability required for real-world clinical deployment (Hua et al., 2025a; Hua et al., 2025b). Building a scalable triage system requires moving beyond black-box classification to establish transparent processing of psychiatric data.

Bridging the gap between unstructured EHR narratives and structured triage decisions remains a primary hurdle in computational psychiatry (Garriga et al., 2023). To solve this, the research team developed a framework that transforms clinical notes into a 20-factor patient profile. Drawing on established clinical frameworks, this profile operationalizes 17 background domains spanning diagnostic, behavioural, functional, and psychosocial presentation, alongside three treatment-action factors. We deployed an LLM to extract these factors from a large-scale Mayo Clinic repository. The model then used the 17 background factors to predict severity labels derived independently from the treatment actions (Arowosegbe & Oyelade, 2023). This architecture separates factor extraction and severity labelling onto non-overlapping feature sets, ensuring predictions are grounded in background clinical presentation rather than the treatment actions used as reference labels. This investigation evaluated whether factor weights autonomously derived by the LLM from patient data provide a significant and replicable performance gain over randomized and unweighted baselines, and whether the structure the LLM independently identified aligns with established clinical determinants of psychiatric severity.

## Methodology

### Computational Infrastructure

All computational tasks, including data preprocessing, natural language extraction, and severity prediction, were executed using Python 3.12 with *pandas*, *SciPy*, and *statsmodels*. To manage the scale of the dataset and run the LLMs in parallel, a cluster of 20 NVIDIA A100 Tensor Core GPUs was used (∼75 hours of use). This infrastructure supported efficient batch processing and prevented computational bottlenecks during the repeated resampling and evaluation phases.

### Data Source & Cohort Selection

This study used de-identified EHR data obtained from the Mayo Clinic *Platform_Accelerate* environment, encompassing over 2.7 million unique patients across Mayo Clinic sites in the United States over the past five years. The dataset included both structured data (e.g., demographics, vitals) and unstructured narrative documentation (e.g., admission notes, discharge summaries, psychiatric evaluations). To isolate clinical encounters containing meaningful psychiatric content, a multi-stage filtering pipeline was developed (Figure 1).

**Figure 1.**
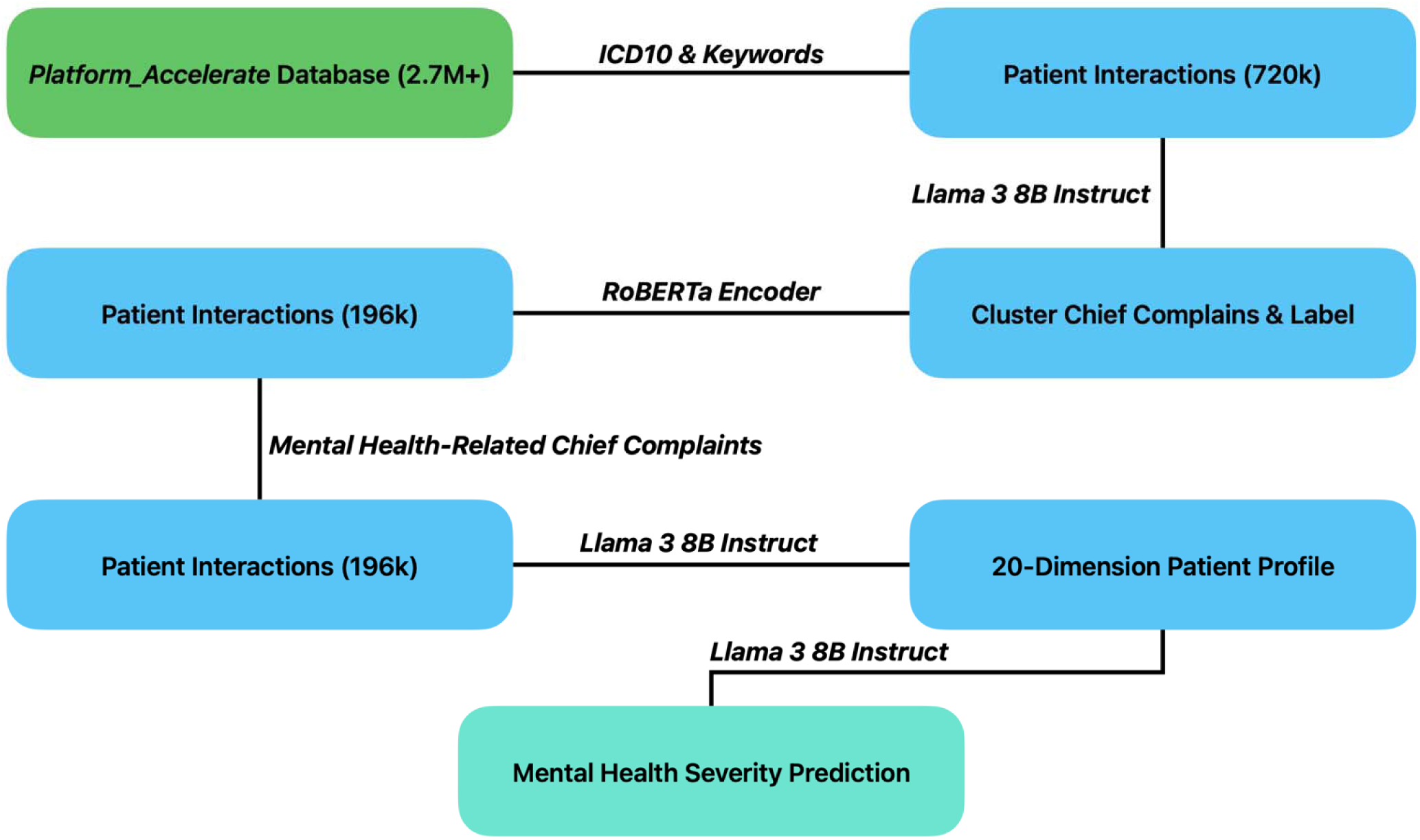
Data reduction and processing pipeline from 2.7M+ encounters to the analytic cohorts.

First, data were filtered for encounters associated with mental health-related ICD-10 codes (e.g., ICD-10 F10-19, F20-29, F30-39, etc.) and keyword matches (e.g., “suicidal ideation”, “anxiety”, “depression”) within the clinical text. Following this, a string-matching algorithm was used to isolate the initial segment of each note containing the reason for referral and Llama 3 8B Instruct was employed to extract the specific chief complaint and reason for referral for each interaction. To exclude irrelevant visits (e.g., routine wellness checks or non-psychiatric somatic complaints), a RoBERTa-based embedding algorithm (princeton-nlp/sup-simcse-roberta-large) was applied to embed the extracted chief complaints. A K-Means clustering algorithm partitioned these embeddings into 100 distinct topic clusters. From each cluster, 100 samples were reviewed by the research team to assign a dominant semantic theme (e.g., “abdominal pain,” “anxiety”, “joint pain”). Clusters without a clear common theme (∼10%) or with non-psychiatric labels were discarded. Only clusters explicitly related to mental health themes (∼25% of the total) were retained. Finally, templated or administrative notes were removed for final filtering to ensure sufficient narrative density was present for factor extraction.

This process yielded ∼196,000 encounters suitable for analysis. From this pool, a 5,000-patient discovery cohort (5k), used for LLM-driven factor-weight derivation and within-sample performance confirmation, and a 10,000-patient replication cohort (10k), used as the primary independent test of whether the derived weight structure generalizes to new data, were sampled. Sampling was performed at the encounter level; no patient-level aggregation was applied.

### Structured Clinical Factor Development

A structured factor-based framework was developed with an expert psychiatrist to operationalize clinical decision-making. Twenty core factors essential for diagnosis and treatment decision-making were identified, drawing on established clinical frameworks for psychiatric assessment. The LLM was used both to extract these factors from patient notes and, in a subsequent step, to autonomously derive predictive weights for each factor from patient data. These factors spanned socio-economic background, demographics, trauma history, substance use, physical comorbidities, social support, and safety concerns. Using a one-shot learning approach, an LLM (Llama 3 8B Instruct) extracted answers to these 20 questions directly from the unstructured clinical notes. These factors were categorized into (1) background (17 high-level diagnostic, behavioural, functional, and psychosocial domains, each comprising multiple specific variables describing the patient’s presentation) and (2) treatment/outcome (3 factors: clinician-documented actions and treatment plans, medication management, and crisis/safety planning).

### Severity Labeling

To establish a reference standard for triage, the intensity of clinical intervention was used as a proxy for severity (intervention-driven severity). Using an LLM algorithm (Llama 3 8B Instruct), treatment/outcome factors (3-factors) were analyzed to assign a binary severity label (severe vs. non-severe) based on pre-specified clinical criteria. Severe criteria included hospital admission (voluntary/involuntary), immediate emergent evaluation, close precautions for suicide/self-harm, use of physical restraints, administration of intramuscular antipsychotics, prescription of multiple simultaneous psychotropics for acute control, or documented lack of response to prior treatment. Non-severe criteria included cases manageable with routine outpatient care or those requiring urgent but non-emergent follow-up. Although both labelling and prediction used LLMs, the models operated on non-overlapping feature sets to reduce information leakage.

Before full-scale deployment, an expert member of the research team conducted a manual validation of the LLM-derived severity labels. A random sample of 100+ de-identified patient notes was independently reviewed, with severity classified as severe or non-severe based solely on the three treatment-action factors. Compared to the LLM-derived labels, this yielded 94.00% accuracy, 100.00% precision, and 83.33% recall, indicating that the automated labelling procedure closely approximated human clinical judgement and that false positives were effectively absent.

### Predictive Weighting & Model Architecture

The core analysis evaluated whether the 17 background factors could accurately predict the treatment-derived severity label. This required two distinct procedures, each with its own batch parameters: factor-weight discovery and severity-prediction evaluation. For factor-weight discovery, the extracted background factors and severity labels were fed into the LLM in 50-patient batches, with the model instructed to extract the most important factors for predicting severity and to assign them numerical weights. This included parallel extraction for factors predictive of severe outcomes and factors indicative of non-severe or protective outcomes. The discovery loop was repeated across 100 iterations per cohort to generate a stable set of weights.

Factor-weight dictionaries from these 100 iterations were then aggregated using a second Llama 3 8B Instruct summarization step. The per-batch dictionaries were concatenated and passed to the LLM with instructions to consolidate overlapping terms (e.g., “suicidal ideation” and “SI”) and average the numerical weights for each consolidated factor. No minimum frequency threshold was applied during this consolidation, yielding a single aggregated dictionary per cohort.

To ensure predictive validity and prevent data leakage, the pipeline used a train-test design. The 5k cohort served as the discovery set: the LLM analyzed EHR notes to derive factor-weight dictionaries, which were then fixed as static inputs for evaluation. The 10k replication cohort had the same pipeline applied independently using the derived dictionaries, with no access to the 5k data. Performance on the 10k cohort, therefore, reflects genuine generalization to unseen data rather than within-sample consistency. Within each cohort, information leakage was further prevented by extracting severity labels and severity predictors from non-overlapping sections of each patient note.

### Experimental Design

To evaluate whether the LLM-derived factor weights could independently predict severity, the pipeline was applied in its evaluation configuration: factor-driven severity prediction was compared against intervention-driven severity labels as the reference standard. Severity prediction from the background factors was operationalized as a weighted composite. The study design was explicitly structured to test three primary hypotheses: (1) that LLM-derived structured weighting schemes would outperform ablated or unweighted baselines; (2) that performance would remain stable across independent sampling iterations; and (3) that the highest-weighted factors would align with established clinical indicators (Zimmerman et al., 2018). As LLMs can generate plausible outputs even in the absence of relevant evidence, multiple control conditions were incorporated to ensure that severity prediction depended on structured clinical information rather than model priors or uncontrolled generative behaviour (Esmaeilzadeh, 2025). The following five weighting schemes were evaluated:

1. *Severe_factors*: A structured weight dictionary autonomously derived by the LLM from the 5k discovery cohort, capturing variables the LLM identified as predictive of severe psychiatric presentations. Fifteen risk-oriented variables received weights between 0.1 and 0.8, including: suicidal ideation, history of trauma, substance use disorder, history of self-injurious behaviour, impulsivity, hopelessness, lack of adherence to treatment recommendations, poor treatment access, environmental risk factors, lack of social supports, chronic pain, chronic stress, history of depression, history of anxiety, and history of PTSD.
2. *Mild_factors*: A parallel LLM-derived dictionary capturing variables the LLM identified as predictive of mild or protective presentations. Eight protective variables received weights between 0.1 and 0.8 (social support, good coping skills, positive social support, regular exercise, healthy diet, good sleep hygiene, routine, and healthy stress management).
3. *Random_weights*: The identical variables from the *Severe_factors* condition, but with randomly generated weights between 0.1 and 0.8. This condition removes the weight signal while preserving the LLM-derived variable selection.
4. *Random_factors*: Nine variables with no clinical relevance to psychiatric severity (e.g., walking, blood type, eye colour) assigned arbitrary weights. This condition removes both variable selection and meaningful weighting.
5. *No_factors*: The LLM was asked to classify severity directly from the patient summary without any structured variables or weights. This condition serves as a zero-shot baseline representing the model’s default predictive capacity (Schnepper et al., 2025)

### Evaluation Procedure

For severity-prediction evaluation, each cohort was assessed across 100 target iterations per scheme. In each iteration, a random subsample of 50 patient notes was drawn without replacement from the cohort pool; no cross-iteration exclusion was applied, so the same patient could appear in multiple iterations, consistent with a bootstrapping evaluation design. The achieved counts fell slightly below the target (∼91–98 iterations for 5k; ∼183–189 for 10k) due to a small number of LLM timeouts and unparseable responses that were silently skipped by the pipeline’s error handling. The Llama 3 8B Instruct model predicted severity using the specified factor weights, and predictions were compared against the intervention-driven reference labels. Performance metrics were averaged across iterations. Across all weighting schemes, this yielded 928 valid evaluations in the 10k replication cohort and 478 in the 5k discovery cohort, totalling 1,406.

### Statistical Analysis

To test whether weighting schemes produced statistically significant differences, *No_factors* served as the primary comparator. Welch’s t-tests were used for pairwise comparisons, Mann-Whitney U tests were used when assumptions were violated, and one-way ANOVA tests were used for comparisons across all schemes. Levene’s tests indicated unequal variances across schemes (accuracy W=3.973, p=0.003; precision W=5.359, p<0.001; recall W=3.973, p=0.003), making Tukey’s HSD inappropriate. Games-Howell post-hoc tests were therefore applied for the primary replication cohort. For the discovery cohort, Levene’s test confirmed equal variances across schemes (accuracy W=0.884, p=0.473; precision W=0.820, p=0.513; recall W=0.884, p=0.473), and Tukey’s HSD was applied instead. To evaluate demographic effects, demographic associations were assessed using Spearman correlations at the iteration level within each cohort. A supplementary pooled regression across both cohorts (n=1,406) confirmed that neither mean age nor female count was an independent predictor of accuracy after adjusting for the weighting scheme. All analyses were conducted using Python 3.12 with *pandas*, *SciPy*, and *statsmodels*. Statistical significance was defined as p<0.05. Under the class-weighted averaging configuration used, accuracy and weighted recall are identical for binary classification (both reported in Table 2 for completeness). As iterations involved resampling from the same underlying cohorts, performance statistics reflect stability and comparative differences rather than independent population estimates.

### Ethical Considerations

All data followed strict de-identification policies governed by Mayo Clinic’s *Platform_Accelerate* data use agreement. A minimum cell size of 10 patients was required for any exported aggregate, meaning individual-level records were not exported. As a consequence, demographic variables (age, sex) were only available as cohort-level summary statistics, averaged across patients within a given folder and dataset combination. These iteration-averaged demographic summaries are therefore the same across all iterations within a scheme, which has implications for the interpretation of demographic analyses: iteration-level correlations between demographic variables and accuracy reflect between-scheme differences in cohort composition rather than genuine patient-level demographic effects on model performance. All analyses were conducted on aggregated data consistent with these export constraints.

## Results

### Dataset

A total of 15,000 de-identified patient notes were analyzed across a 5k discovery cohort and a 10k replication cohort, randomly sampled from the filtered pool of ∼196,000 eligible encounters identified through the 2.7 million record preprocessing pipeline. Each record contained rich narrative documentation of psychiatric symptoms, psychosocial context, and treatment planning.

Demographic characteristics were consistent across weighting conditions within each cohort (Table 1). In the replication cohort (10k), mean age ranged from 42.07 to 42.16 years, and the proportion of female patients averaged 69.04%. In the discovery cohort (5k), mean age ranged from 40.65 to 40.81 years, and the proportion of female patients averaged 69.00%. The proportion of patients classified as clinically severe based on the LLM-derived treatment labels ranged from 18.96% to 51.04% across schemes in the replication cohort and from 24.49-47.02% in the 5k cohort, reflecting varying emphasis of each weighting condition rather than true cohort differences in clinical severity.

**Table 1.** Demographic composition and severity distribution across weighting schemes for the primary replication cohort (10k). Values represent iteration-averaged estimates derived from de-identified patient notes exported under Mayo Clinic’s *Platform_Accelerate* minimum cell-size constraints (≥10 patients per cohort). Differences in reported severity prevalence across conditions reflect the varying emphasis of each weighting scheme rather than true cohort differences in clinical severity; all conditions were evaluated on the same underlying replication cohort.

| Weighting Scheme | Total N (Female) | Mean Age | Severe N (%) |
| --- | --- | --- | --- |
| *Severe_factors* | 9,089 (6,273) | 42.07 | 3,772 (41.50) |
| *Mild_factors* | 8,534 (5,896) | 42.10 | 3,625 (42.48) |
| *Random_weights* | 8,534 (5,896) | 42.10 | 4,356 (51.04) |
| *Random_factors* | 8,763 (6,044) | 42.16 | 1,661 (18.95) |
| *No_factors* | 8,811 (6,082) | 42.15 | 3,032 (34.41) |

Welch’s t-test confirmed no statistically significant differences in aggregate accuracy (p=0.829) or recall (p=0.829), with a marginal difference in precision (p=0.084). Consistent with the study design, primary results are reported from the 10k replication cohort; 5k discovery cohort results are provided as within-sample confirmation.

### LLM-Derived Factor Weights

Before the evaluation phase, the LLM was applied to the 5k discovery cohort to identify which background factors and variables best predicted psychiatric severity and to assign numerical weights reflecting each variable’s relative importance. Across 100 iterations of 50-patient batches, the model consistently converged on a set of risk-oriented variables as the strongest predictors of severe presentations. Following aggregation and consolidation of overlapping terms across iterations, the resulting *Severe_factors* dictionary assigned the highest weights to suicidal ideation (0.8), history of trauma (0.7), substance use disorder (0.6), history of self-injurious behaviour (0.5), and impulsivity (0.5), with ten additional variables weighted between 0.1 and 0.4. A parallel extraction targeting mild or protective presentations produced the *Mild_factors* dictionary, with social support (0.8), good coping skills (0.7), and positive social support (0.6) receiving the highest weights, and five additional protective variables weighted between 0.1 and 0.5 (Figure 4). Notably, the variables and weights the LLM independently derived closely mirror established clinical determinants of psychiatric severity.

### Model Performance Across Weighting Schemes

In the primary replication cohort (10k), the LLM successfully predicted mental health severity from the 17 background factors, but performance varied significantly depending on the weighting scheme applied. A one-way ANOVA confirmed this variance was highly significant, F(4, 923)=11.419, p<0.001, indicating that the design of factor weighting directly influences predictive accuracy. The *No_factors* condition served as the foundational baseline, establishing the model’s default predictive capacity (accuracy=0.577, SEM=0.016).

LLM-derived structured schemes (*Severe_factors*, *Mild_factors*) significantly outperformed the baseline. *Mild_factors* achieved the highest mean accuracy at 0.717 (SEM=0.018), with *Severe_factors* closely equivalent at 0.711 (SEM=0.017). *Random_weights*, which retained the LLM-derived variable selection but randomized the weights, yielded an accuracy of 0.665 (SEM=0.019), while *Random_factors*, which used clinically irrelevant variables, yielded an accuracy of 0.660 (SEM=0.013). Changes in the weighting scheme affected the precision and recall equally across conditions (Figure 2; Figure 3), indicating that structured weighting improved overall discriminative capacity rather than shifting the model’s classification threshold.

**Figure 2.**
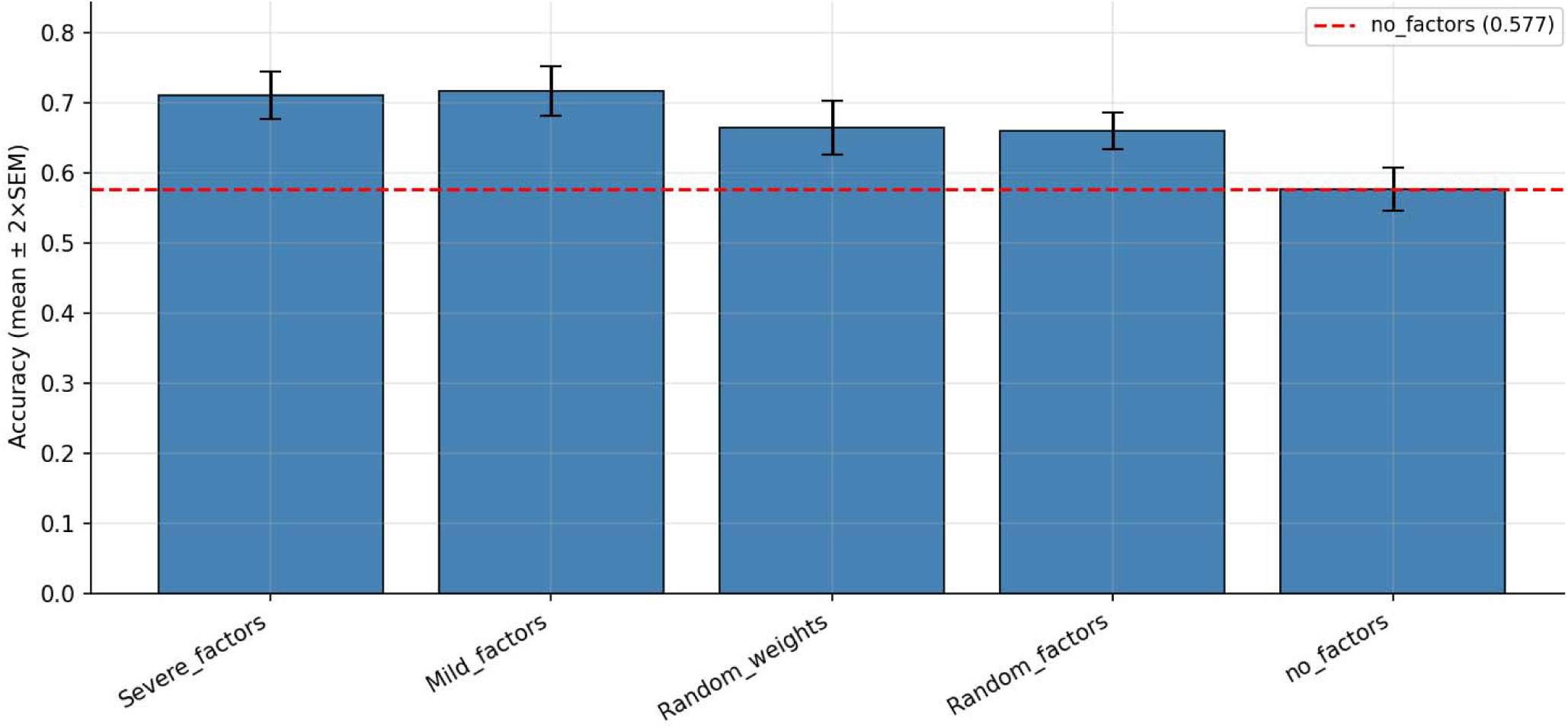
Model accuracy (mean ± 2×SEM) for each weighting scheme in the primary replication cohort (10k, n=928 iterations). A dashed horizontal line indicates the *No_factors* zero-shot baseline (accuracy=0.577).

**Figure 3.**
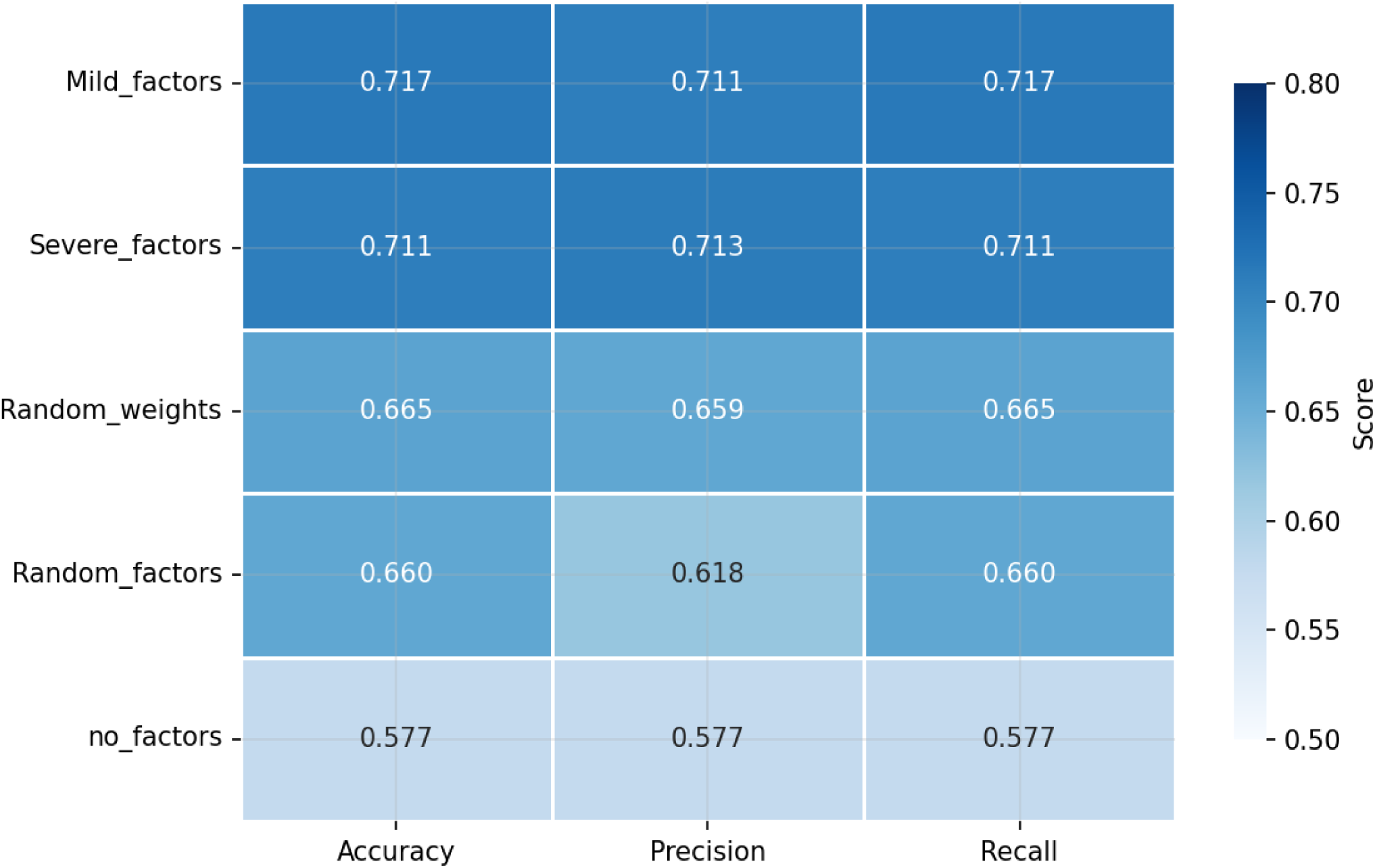
Model performance heatmap (accuracy, precision, and recall) for each weighting scheme in the primary replication cohort (10k). LLM-derived structured conditions (*Mild_factors*, *Severe_factors*) achieved substantially higher scores across all three metrics than the controls and the zero-shot baseline.

**Figure 4.**
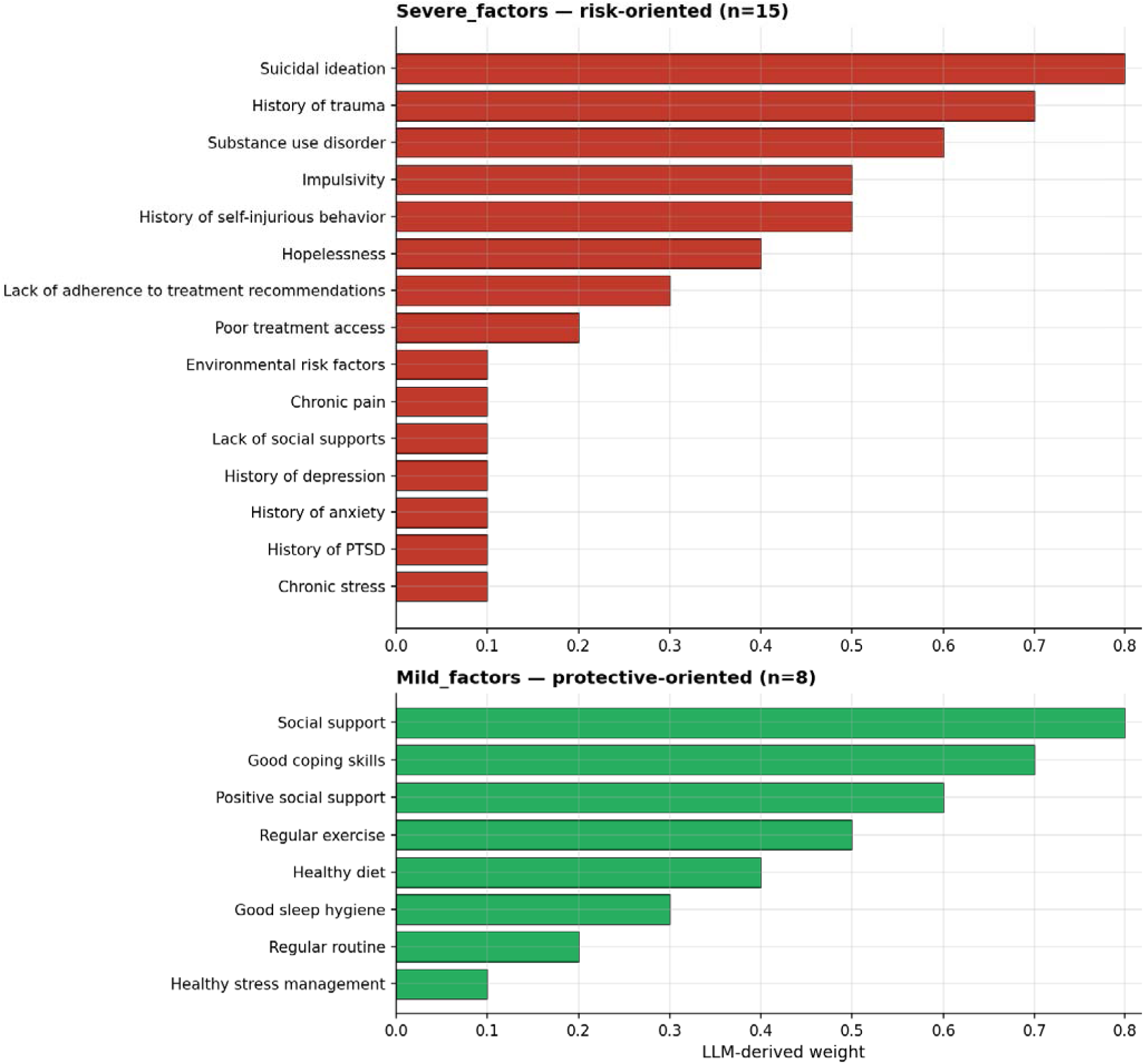
LLM-derived weight dictionaries for the two structured weighting schemes, generated by prompting Llama 3 8B Instruct to identify predictors of psychiatric severity from 5k discovery cohort patient notes and to assign numerical weights, averaged across 100 iterations. (Top) *Severe_factors* 15 risk-oriented variables. (Bottom) *Mild_factors*: 8 protective variables. Despite operating on non-overlapping variable sets, both conditions achieved statistically equivalent predictive accuracy in the replication cohort (*Severe_factors*=0.711, *Mild_factors*=0.717; Games-Howell p=0.999), and both significantly outperformed the baseline (*No_factors*=0.577; both p<0.001).

Within-sample confirmation on the 5k discovery cohort showed similar results to the replication cohort, with *Severe_factors* achieving an accuracy of 0.724 (SEM=0.022), followed by *Random_weights* (0.691), *Mild_factors* (0.679), *Random_factors* (0.622), and *No_factors* (0.592). The slight elevation of *Severe_factors* relative to *Mild_factors* in the 5k cohort is consistent with a within-sample advantage. The *Severe_factors* dictionary was derived from the same 5k data. The convergence of LLM-derived schemes to equivalent performance in the 10k cohort confirms that this advantage does not reflect the genuine superiority of one scheme over another.

### Comparison to Baselines

Pairwise comparisons against *No_factors* demonstrated consistent and significant improvements for all structured conditions (Table 2). LLM-derived schemes were significantly more accurate than baseline (*Mild_factors* t=5.906, p<0.001, Hedges’ g=0.612; *Severe_factors* t=5.827, p<0.001, Hedges’ g=0.598). The control conditions also significantly outperformed baseline (*Random_weights* and *Random_factors* p<0.001), though with smaller effect sizes (*Random_weights* Hedges’ g=0.368, *Random_factors* Hedges’ g=0.418).

**Table 2.** Mean performance metrics for each LLM-derived and control weighting scheme in the primary replication cohort (10k). Differences (Δ) represent the absolute change in performance relative to the *No_factors* zero-shot baseline (accuracy=0.577, precision=0.577, recall=0.577). Effect sizes were calculated as Hedges’ g alongside their corresponding p-values (Welch’s t-test vs No_factors). Dashes indicate the baseline condition, for which no comparative effect size is applicable.

| Weighting Scheme | Accuracy |  | Precision |  | Recall |  |
| --- | --- | --- | --- | --- | --- | --- |
| | Mean ($\Delta$) | Hedges' g (p) | Mean ($\Delta$) | Hedges' g (p) | Mean ($\Delta$) | Hedges' g (p) |
| *Mild_factors* | 0.717 (0.140) | 0.612 (<0.001) | 0.711 (0.134) | 0.502 (<0.001) | 0.717 (0.140) | 0.612 (<0.001) |
| *Severe_factors* | 0.711 (0.134) | 0.598 (<0.001) | 0.713 (0.136) | 0.514 (<0.001) | 0.711 (0.134) | 0.598 (<0.001) |
| *Random_weights* | 0.665 (0.088) | 0.368 (<0.001) | 0.659 (0.083) | 0.283 (0.007) | 0.665 (0.088) | 0.368 (<0.001) |
| *Random_factors* | 0.660 (0.083) | 0.418 (<0.001) | 0.618 (0.041) | 0.204 (0.049) | 0.660 (0.083) | 0.418 (<0.001) |
| *No_factors* | 0.577 |  | 0.577 |  | 0.577 |  |

Post-hoc Games-Howell tests further confirmed that pairwise differences between *Mild_factors* and *Severe_factors* were insignificant (p=0.999), and both significantly outperformed all control conditions (all p<0.004). This indicates that once a coherent, LLM-generated factor structure is imposed, the specific weighting design is less consequential than the presence of structure itself.

### Precision-Recall Trade-off

Precision-recall analysis showed consistent differentiation among weighting schemes (Figure 3). LLM-derived structured schemes achieved strong and balanced performance (*Mild_factors* precision=0.711, recall=0.717; *Severe_factors* precision=0.713, recall=0.711) compared to the *No_factors* baseline (precision=0.577, recall=0.577). A strong positive correlation between precision and recall (Spearman ρ=0.819, p<0.001) suggested that weighting scheme differences reflected changes in discriminative capacity rather than shifts in classification threshold. This pattern was held in the 5k confirmation cohort (Spearman ρ=0.823, p<0.001), indicating that the precision-recall relationship is stable across independent samples.

### Demographic Associations

Demographic composition was consistent across weighting conditions within each cohort (Table 1), with mean age varying by less than 0.09 years and female proportion by less than 0.12 percentage points across schemes. Iteration-level Spearman correlations revealed statistically significant but small associations between mean age and accuracy (ρ=-0.272, p<0.001), and between sex distribution and accuracy (Male_Count ρ=-0.092, p=0.005, Female_Count ρ=-0.092, p=0.005). Given that demographic values are scheme-level constants within each cohort, varying by at most 0.09 years across schemes, these iteration-level correlations reflect between-scheme demographic differences rather than genuine patient-level demographic effects on model performance. A supplementary pooled regression across both cohorts (n=1,406), which introduced meaningful demographic variation between the 5k and 10k cohorts, confirmed that neither mean age nor female count was an independent predictor of accuracy after adjusting for the weighting scheme (Mean_Age p=0.419; Female_Count p=0.427; F(6, 1,399)=10.26, p<0.001). The overall model effect was driven by the weighting scheme covariate. These results suggest that observed accuracy differences were driven by algorithmic weighting configurations rather than differences in sample composition or demographic case mix.

### Severity-Accuracy Association

We tested whether variation in severe-case prevalence across iteration subsamples was associated with model accuracy. A Spearman correlation revealed a statistically significant but weak positive association (Spearman ρ=0.235, p<0.001, R^2^=0.056). This association most likely reflects a base-rate sampling artifact: iterations with a higher proportion of severe cases may contain more clinically distinctive presentations, marginally inflating raw accuracy independent of true model quality. The effect size (accounting for ∼5.5% of iteration-level variance in accuracy) suggests this has minimal practical impact on the primary conclusions. Precision and recall were computed with class-weighted averaging and are less susceptible to this influence, and showed consistent patterns across all comparisons.

## Discussion

This study demonstrates that an LLM can derive clinically meaningful factor weights from unstructured psychiatric EHR narratives and use those self-derived weights to predict clinician-implied severity at scale. By separating severity labelling, derived from three treatment-action factors, from severity prediction, based on 17 background narrative factors, the study provides direct evidence that an LLM can replicate the severity judgements implied by clinician treatment decisions using only narrative EHR content, without manual annotation or pre-specified clinical rules.

A particularly notable finding was the nature of the factor structure that the LLM independently derived from patient narratives. Without being told which clinical variables matter, the model converged on suicidal ideation, history of trauma, substance use disorder, and history of self-injurious behaviour as the highest-weighted predictors of severe psychiatric presentations. These are variables that psychiatric research and validated assessment instruments, including the Columbia Suicide Severity Rating Scale, the PHQ-9, and established risk stratification frameworks, have identified as the core determinants of psychiatric severity and acute risk (Kroenke et al., 2001; Posner et al., 2011; Zimmerman et al., 2018). The LLM arrived at this structure by analyzing unstructured patient notes, with no exposure to the clinical literature and no guidance about which variables to prioritize. That a general-purpose language model, prompted only to identify predictors of severity from EHR text, independently recovered the same factor structure that clinicians and researchers have constructed is notable. This speaks to the validity of the approach and the degree to which the LLM’s high-weight factors align with established clinical determinants.

Across the replication cohort, the *No_factors* condition established the zero-shot baseline, reflecting the model’s default predictive capacity in the complete absence of structured input (accuracy=0.577). Against this baseline, the value of the LLM-derived factor structure becomes clear. LLM-derived structured schemes (*Mild_factors*=0.717; *Severe_factors*=0.711) achieved substantial improvements over the baseline, while the controls performed intermediately (*Random_weights*=0.665; *Random_factors*=0.660). This hierarchy confirms that performance gains were driven by the structured factor space rather than intrinsic LLM priors or uncontrolled generative behaviour (Esmaeilzadeh, 2025). These results replicated the pattern observed in the discovery cohort, confirming the stability and generalizability of the findings across independent samples.

The performance gradient across conditions reveals what each element of the LLM-derived structure contributes. *Random_factors*, which used clinically irrelevant variables with arbitrary weights, performed only marginally above the baseline, suggesting that structure alone, without meaningful content, provides limited benefit. *Random_weights*, which retained the LLM-derived variable selection but randomized the weights, performed similarly to *Random_factors*, indicating that in the absence of meaningful weights, even clinically relevant variable selection adds little. The jump to the LLM-derived structured schemes demonstrated that both the variable selection and the weight values the LLM assigned carry a genuine predictive signal. The LLM did not simply benefit from being given any structure; it benefited from the structure it had derived from patient data.

Notably, *Mild_factors* and *Severe_factors* produced statistically equivalent accuracy despite using opposing clinical constructs, one capturing risk indicators of severe presentations, the other capturing protective indicators of mild presentations. The convergence to equivalent performance suggests that the LLM responds to the coherence and informativeness of a factor structure rather than to its specific clinical direction. Whether the structure encodes risk or protection, the presence of a meaningful weighting scheme is sufficient to improve severity prediction over unstructured baselines.

These findings support the central premise that scalable severity prediction from EHR narratives is feasible without human annotation, standardized assessments, or direct expert clinician involvement at the point of triage (Qventus, 2026). Treatment actions documented in the EHR provide a clinically grounded reference standard, and the LLM’s ability to map background narrative features to those treatment-derived outcomes demonstrates a viable path toward automated severity estimation across large patient populations. This scalability benefit is particularly relevant given that the analytic cohorts were drawn from a preprocessing pipeline spanning more than 2.7 million encounters, suggesting the approach is applicable at the health-system scale.

### Implications

The above findings have direct implications for clinical practice and methodological development. From a clinical perspective, these results highlight the potential of LLM-driven factor extraction to support real-time triage, risk monitoring, and population-level surveillance, supporting integration within clinical workflows and retrospective analysis of large-scale EHR datasets to identify trends, unmet needs, or treatment gaps across health systems. Because the severity labels were derived from clinician decision-making, hospital admission, crisis precautions, and antipsychotic administration, the system captures how clinicians act on severity rather than simply how symptoms are described (Zimmerman et al., 2018). This distinction is particularly relevant for triage, where decisions hinge on anticipated resource needs rather than symptom counts alone, and supports prospective integration within clinical decision support workflows.

Methodologically, the study introduces an interpretable and modular framework for evaluating LLM performance in psychiatric classification. Rather than relying on opaque end-to-end predictions, the approach uses explicit intermediate representations, 17 background factors with associated weights, and systematically tests how manipulating those weights affects performance. The control design, in which variable selection and weight values are independently varied across conditions, disentangles the contribution of each element of the LLM-derived structure. The consistent performance across hundreds of independent subsampling iterations, with moderate within-scheme variability, indicates that the mapping between background narrative factors and treatment-derived severity labels is stable across patient samples. This stability supports the potential use of this framework in clinical decision support systems, digital phenotyping applications, and population-level mental health surveillance.

### LLMs vs Traditional Regression Models

While traditional machine learning and regression models are standard tools for prediction, their utility in mental health is often bottlenecked by data format. Logistic regression and random forests require highly structured, tabular data. In real-world psychiatric workflows, extracting this structured data from narrative text requires extensive manual chart review or the use of rigid, keyword-based natural language processing (NLP) pipelines that miss contextual nuance. The LLM framework deployed here circumvents this limitation by directly ingesting unstructured clinical narratives and autonomously structuring the 17 background factors.

It is crucial to recognize, however, that LLMs are not statistical regression models. When an LLM is prompted to predict or apply specific factor weights, it relies on semantic approximation rather than exact mathematical calculation. Because we cannot guarantee precise adherence to applied weights, pure predictive performance might actually be higher if a traditional regression model were used on pre-structured data. LLMs operate through an inherent interpretation layer that complicates standard mathematical validation. This unavoidable interpretation layer, along with computational expense, hallucination risk, and non-deterministic outputs, highlights the distinct limitations of LLMs. Traditional regression models, once trained, are deterministic, lightweight, and mathematically transparent. Therefore, rather than replacing regression models, LLMs are best suited as an upstream extraction layer where they can convert messy clinical narratives into the structured, quantifiable factors that traditional predictive models or clinical decision support rules require.

### Interpretability

A major advantage of this factor-based approach is its interpretability. By decomposing predictions into distinct conceptual domains, the model allows users to understand not only whether a prediction is severe but also why. The factor decomposition is transparent by design: users can inspect which variables drove a given prediction and at what weight. The variables the LLM assigned the highest weights, suicidal ideation, history of trauma, and substance use disorder, are the same domains clinicians consult when assessing acute risk, meaning the model’s reasoning is legible to clinical practitioners without specialist ML knowledge (Kroenke et al., 2001; Posner et al., 2011; Spitzer et al., 2006). This correspondence, which emerged from the LLM’s analysis of patient data rather than from pre-specification, suggests that LLMs can operationalize the same semantic cues clinicians rely upon when making severity and risk-related decisions, and that this operationalization can be surfaced through structured prompting to recover treatment-derived severity from narrative text.

The observed precision-recall improvements demonstrate that structured weighting amplified meaningful signals derived from the background factors rather than inducing threshold shifts or overfitting. These findings align with emerging work showing that structured prompts and domain-informed embeddings reduce semantic drift and improve interpretability in clinical LLM applications (Sonoda et al., 2025).

### Comparison with Prior Work

Previous studies have shown that LLMs can classify psychiatric phenotypes from text, but most treat the problem as a black-box task with limited interpretability (Flathers et al., 2025). This study extends that line of work by introducing a transparent, factorized architecture that enables direct hypothesis testing about which clinical domains contribute most strongly to severity prediction.

To the authors’ knowledge, this is the first large-scale demonstration of a single-LLM severity framework in which the same model derives predictive factor weights from patient data and subsequently applies those weights to predict treatment-derived severity labels across a replication cohort. This architecture reduces reliance on manually annotated datasets by using treatment-derived labels as the reference standard and could enable sustainable and scalable solutions for real-world applications. Clinician behaviours can encode biases, which must be recognized. The strong convergence between the LLM-derived factor weights and established clinical theory supports the clinical plausibility of the approach and suggests that the findings are not an artifact of the specific patient sample or institutional context.

### Limitations

Several limitations merit consideration. First, the data were drawn from a single health system, which may limit generalizability to institutions with different documentation styles or clinical practices. Future validation across multi-institutional datasets is warranted. Second, the severity labels were derived from clinician behaviour rather than formal diagnostic criteria, meaning the LLM may inherit documentation or practice-pattern biases. Third, although the labelling and prediction steps operate on non-overlapping feature sets within each note, the same underlying model is used for factor extraction, label derivation, and severity prediction. Shared model representations could, in principle, introduce subtle information leakage that feature-level separation does not detect; future work should validate the framework with separately-trained or architecturally distinct models for labelling and prediction. Fourth, the weighting schemes were evaluated in static form rather than optimized via direct training or reinforcement learning; future work could explore dynamic or learned weighting. The restriction to 20 pre-selected factors means that predictive signals from factors outside this framework (e.g., granular social determinants or specific biomarker data) may have been overlooked. Fifth, the severity labels were based on individual clinician decisions, which may introduce inter-provider variability. Finally, because the single-model architecture produces severity labels without human supervision, inter-rater reliability cannot be directly calculated, although iteration-level metrics confirm high internal stability.

### Future Directions

Future research should extend this framework to longitudinal severity modelling, examining how LLM-derived trajectories correlate with relapse, hospitalization, and treatment outcomes. Incorporating additional data sources (e.g., structured fields, lab values, or prior encounters) may further strengthen predictions. Comparative testing across different LLM families can identify architecture-specific versus architecture-agnostic components of severity inference. Ultimately, embedding this approach in clinical decision support systems could enable real-time, interpretable risk assessment directly from narrative EHR text, supporting more scalable and equitable mental health care.

## Conclusion

This study demonstrates that an LLM can derive clinically meaningful factor weights from unstructured EHR narratives and use them to predict psychiatric severity at scale. By linking background clinical factors to severity labels derived from clinician treatment actions, the proposed framework provides an objective, data-driven approach to estimating urgency and resource needs without reliance on manual annotation or real-time expert review. The results show that while LLMs possess a baseline capacity to infer severity, deriving a structured factor architecture substantially amplifies this signal. LLM-derived factor weighting consistently outperformed both ablated and unweighted baselines, maintaining reproducibility across independent cohorts. The convergence of LLM-derived high-weight predictors with established clinical risk domains supports the face validity and clinical utility of this approach. Ultimately, structured LLM-driven analysis of clinical documentation can complement clinician expertise and support the scalable allocation of care in resource-constrained mental health systems.

## Data Availability

All data produced in the present study are available upon reasonable request to the authors.

## Acknowledgements

The authors wish to thank the Mayo Clinic *Platform_Accelerate* team for their support in providing access to the clinical dataset used in this study.

